# Quantifying Uncertainty in Alzheimer’s Disease Progression Modelling: A Variational Disease Progression Score Framework

**DOI:** 10.64898/2026.08.19.26360855

**Authors:** Tunchanok Ngamsaowaros, Indu Bodala, Sofia Michopoulou, Mahesan Niranjan

**Affiliations:** School of Electronics and Computer Science, University of Southampton, United Kingdom; Nuclear Medicine Physics, University Hospital Southampton NHS Foundation Trust, United Kingdom

## Abstract

Predicting the course of Alzheimer’s disease for individual patients remains a major challenge due to the heterogeneity of disease expression and the sparsity of longitudinal data. We introduce a variational Disease Progression Score (DPS) framework that maps multimodal biomarker dynamics (Cerebrospinal fluid, neuroimaging, and cognitive assessments) onto a continuous latent timeline with quantified uncertainty. The framework combines a neural encoder, which infers subject-specific progression parameters from demographic and clinical features, with a cascade of logistic functions structured according to the amyloid cascade hypothesis. Applied to the Alzheimer’s Disease Neuroimaging Initiative (ADNI) cohort, the inferred timeline separated diagnostic groups it never observed (AUC 0.98 for cognitively normal vs Alzheimer’s Disease), and the estimated cascade strengths and biomarker orderings were consistent with the established sequence of Alzheimer’s pathology. The model produces individualised prognoses for previously unseen subjects from baseline data alone, with 95% credible intervals achieving 89-98% empirical coverage across biomarkers, and these predictions can be dynamically refined as new observations become available. The framework thus provides a biologically interpretable, uncertainty-aware index of disease severity, offering a probabilistic foundation for patient-level prognosis and precision monitoring in Alzheimer’s disease.

**Author summary:** Alzheimer’s disease develops gradually, and its course varies widely from person to person, which makes it hard to predict how any individual will progress. Accurate, individual-level predictions would help doctors intervene earlier and design better clinical trials, but the data available in practice are sparse and collected at irregular times, and most existing prediction tools offer little sense of how confident they are.

We developed a method that places each person on a shared disease timeline estimated from routinely collected information such as age, genetics, and a brief cognitive assessment. Moreover, our model reports the uncertainty of each prediction and refines both the prediction and its uncertainty as new measurements become available for a patient.

Using the ADNI dataset, we found that the model realigned patients according to their clinical diagnoses. It also recovered biological findings consistent with the established understanding of the disease. Its confidence estimates were reliable for most measurements. We hope this approach is a step forward in the development of prediction tools that clinicians can genuinely trust for individual patients.

## Introduction

Alzheimer’s disease (AD) is a progressive neurodegenerative disorder characterised by the accumulation of amyloid-*β* plaques and neurofibrillary tangles, which drive synaptic dysfunction, neuronal loss, and progressive cognitive and functional decline [1]. Dementia affects over 55 million people worldwide, with AD accounting for an estimated 60–70% of cases [2]. In the United Kingdom alone, nearly one million people are affected [3]. Although recent anti-amyloid therapies represent an important advance, their benefit is largely confined to the early stages of the disease, and no disease-modifying cure is currently available for established AD [4, 5]. There is therefore a pressing need for methods that can detect AD early and reliably predict its course at the individual level, supporting timely intervention, stronger clinical trial design, and personalised patient care.

Modelling AD progression and generating individualised predictions is a difficult inference problem because the available clinical data are typically sparse, cross-sectional, and collected asynchronously across patients. The Disease Progression Score (DPS) framework [6–8] addresses this by aligning individuals along a latent disease timeline, under the assumption that biomarker trajectories are monotonically ordered and driven by a shared latent disease state. This yields a common disease scale on which patient-specific trajectories can be derived from population-level observations. However, classical DPS models treat biomarkers independently and so cannot capture inter-biomarker relationships or shared temporal dynamics.

Ordinary differential equation (ODE)-based models address this by describing biomarkers in terms of their rates of change, providing a natural way to represent temporal dependencies and cascade relationships. They can encode established biological hypotheses, such as the amyloid cascade [9, 10], or learn dynamics directly from data without predefined assumptions [11], and have been extended to additional processes such as neural dynamics and molecular-level modulation of progression [12, 13]. However, reliable parameter estimation generally requires dense longitudinal sampling that is rarely available in real-world cohorts.

Mixed-effects models offer an alternative, combining fixed effects that describe population-level patterns with random effects that capture individual deviation. They accommodate linear [14], nonlinear [15], and multivariate [16–18] formulations and handle missing data and irregular follow-up naturally. Random-effect parameters, such as slopes and intercepts, have clear clinical interpretations as individual rates of change and onset times. Their principal weaknesses are restrictive assumptions about trajectory shape (e.g., linear or logistic) and an absence of predictive uncertainty. Bayesian extensions quantify uncertainty at both the population and subject levels [19–21] using Markov Chain Monte Carlo (MCMC) sampling, but are computationally intensive and scale poorly to high-dimensional, multimodal data and large cohorts. Generalisation to unseen subjects is a further obstacle, as predictions for new individuals often require refitting the full model, limiting clinical practicality.

Deep learning approaches, notably recurrent neural network (RNN)-based models [22–25] have also been applied to AD progression for diagnostic prediction and biomarker forecasting. While flexible enough to capture nonlinear, multimodal dynamics, their black-box nature prohibits interpretation and provides limited uncertainty quantification, limiting their reliability in clinical decision-making.

Across these approaches, a common pattern emerges: no existing method simultaneously delivers biological interpretability, calibrated uncertainty quantification, and reliable generalisation to previously unseen patients. We address this gap with a variational extension of the DPS framework. Our central contribution is to couple the interpretability of the DPS formulation and the prior clinical knowledge encoded in the amyloid cascade hypothesis [9] with principled uncertainty quantification through variational inference, improving generalisation both to unseen visits and to previously unseen subjects. We further introduce a dynamic updating mechanism that refines predicted trajectories as new longitudinal observations accrue, mirroring how clinical prognoses are revised over time. Applied to the Alzheimer’s Disease Neuroimaging Initiative (ADNI) cohort [1], the framework produces calibrated, individualised prognoses from baseline data, with estimated cascade strengths and biomarker orderings that are consistent with the established sequence of Alzheimer’s pathology.

## Methods

Building on recent techniques in probabilistic generative modelling in machine learning, we proposed a variational extension of the Disease Progression Score (DPS) framework to jointly model individual disease staging and longitudinal biomarker trajectories. The framework integrates baseline demographic and clinical features with longitudinal measurements to infer each subject’s progression rate and onset time while explicitly quantifying predictive uncertainty. Its overall architecture and workflow are shown in Fig 1.

**Fig 1.**
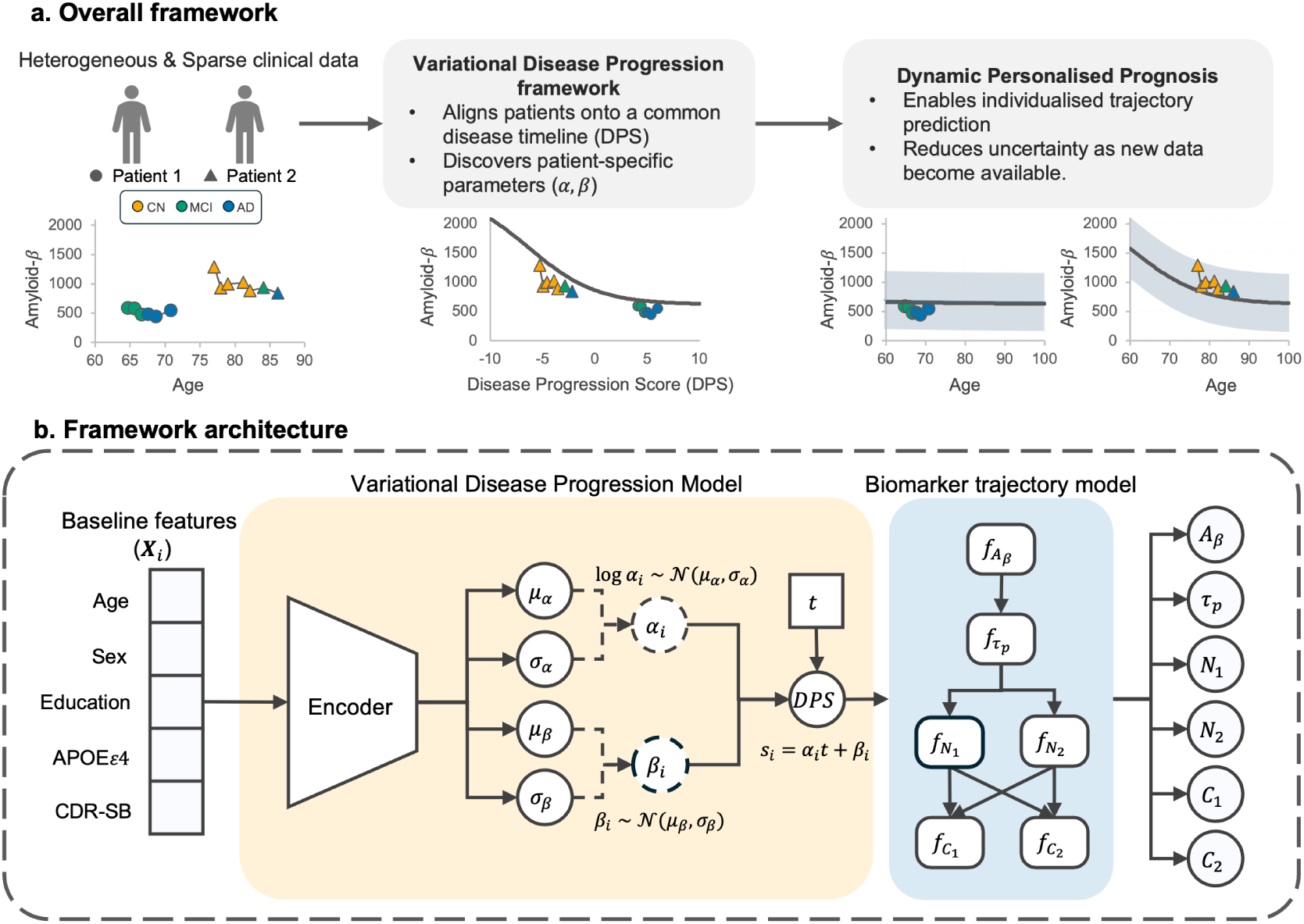
Conceptual and architectural overview of the variational Disease Progression Score (DPS) framework. (a) Overall workflow. The framework takes sparse, asynchronous, multimodal clinical data as input (left), where each point is an observation, marker shape denotes patient identity, and colour denotes diagnostic status. Individuals are mapped onto a continuous latent disease timeline, from which population-level biomarker trajectories are reconstructed (middle). This alignment enables personalised trajectory prediction with explicitly quantified uncertainty that is dynamically refined as new longitudinal observations are incorporated (right). (b) Detailed architecture. For each individual, baseline features (X*_i_*)—age, sex, years of education, APOE*ε*4 genotype, and Clinical Dementia Rating-Sum of Boxes (CDR-SB)—are encoded into the mean (*µ*) and standard deviation (*σ*) of two subject-specific latent parameters: progression rate (*α_i_*) and onset time (*β_i_*). Sampling from these posterior distributions quantifies subject-level uncertainty. The DPS (*s_i_*) is then computed from normalised age (*t*) using these parameters (see the Disease Progression Score (DPS) section). Finally, biomarker trajectories are modelled as cascade functions of the DPS and their parent biomarkers, structured as amyloid-*β →* p-tau *→* medial temporal atrophy (hippocampus, entorhinal cortex) *→* cognitive decline (ADAS-13, CDR-SB), producing predicted biomarker values while capturing hierarchical dependencies between them (see the Biomarker trajectory model section).

The framework consists of two components. In the first, baseline demographic and clinical features—age, sex, years of education, Apolipoprotein E (APOE) genotype, and Clinical Dementia Rating-Sum of Boxes (CDR-SB)—are encoded into posterior probability distributions over two subject-specific latent parameters: progression rate (*α_i_*) and onset time (*β_i_*). These parameters define a continuous Disease Progression Score (DPS) that aligns individuals onto a shared disease timeline. In the second component, each biomarker trajectory is modelled as a cascade function of the DPS and its upstream (parent) biomarkers, following a cascade topology specified a priori from the amyloid cascade hypothesis [9]. This structure represents the hierarchical, temporal ordering of amyloid pathology, tau accumulation, neurodegeneration, and cognitive decline. The framework was trained and evaluated on longitudinal data from the Alzheimer’s Disease Neuroimaging Initiative (ADNI) database [1]. Six key biomarkers were modelled: cerebrospinal fluid (CSF) amyloid-*β* (*Aβ*), CSF phosphorylated Tau (*τ_p_*), MRI-derived hippocampal and entorhinal cortex volumes, the Alzheimer’s Disease Assessment Scale-Cognitive Subscale (13 items, ADAS-13) [26], and the CDR-SB [27].

This formulation supports both population-level trajectory learning and personalised prediction. It can be fine-tuned dynamically. As longitudinal observations become available, the encoder can be fine-tuned to refine these individualised predictions, similar to real-world clinical practice.

### Dataset

The data used for training and evaluation were drawn from the Alzheimer’s Disease Neuroimaging Initiative (ADNI) database [1]. Launched in 2003, ADNI is a large-scale, longitudinal, multicentre study providing publicly available clinical, genetic, imaging, and biochemical biomarker data for Alzheimer’s disease (AD). We used subjects originally enrolled in ADNI-1, together with any follow-up data available from ADNI-GO, ADNI-2, and ADNI-3, retaining those with complete baseline demographic information (age, sex, year of education, and Apolipoprotein E (APOE) genotype).

The ADNI-1 cohort comprises 819 subjects, with baseline and longitudinal characteristics summarised in Table 1. A subject was assigned to the training cohort if they had at least 2 longitudinal measurements for each biomarker or at least 15 observed measurements across all visits. This yielded 611 subjects (74.6% of the total cohort). For each training subject, the last visit was held out as a temporal test set, leaving the earlier visits for training (15,393 measurements across 4,055 visits). The remaining 208 subjects, who did not meet the inclusion criteria, formed a validation cohort of entirely unseen individuals, used to assess subject-level generalisation. As not all biomarkers are recorded at every visit, the number of measurements per biomarker in each split is reported in Table 2.

**Table 1.**
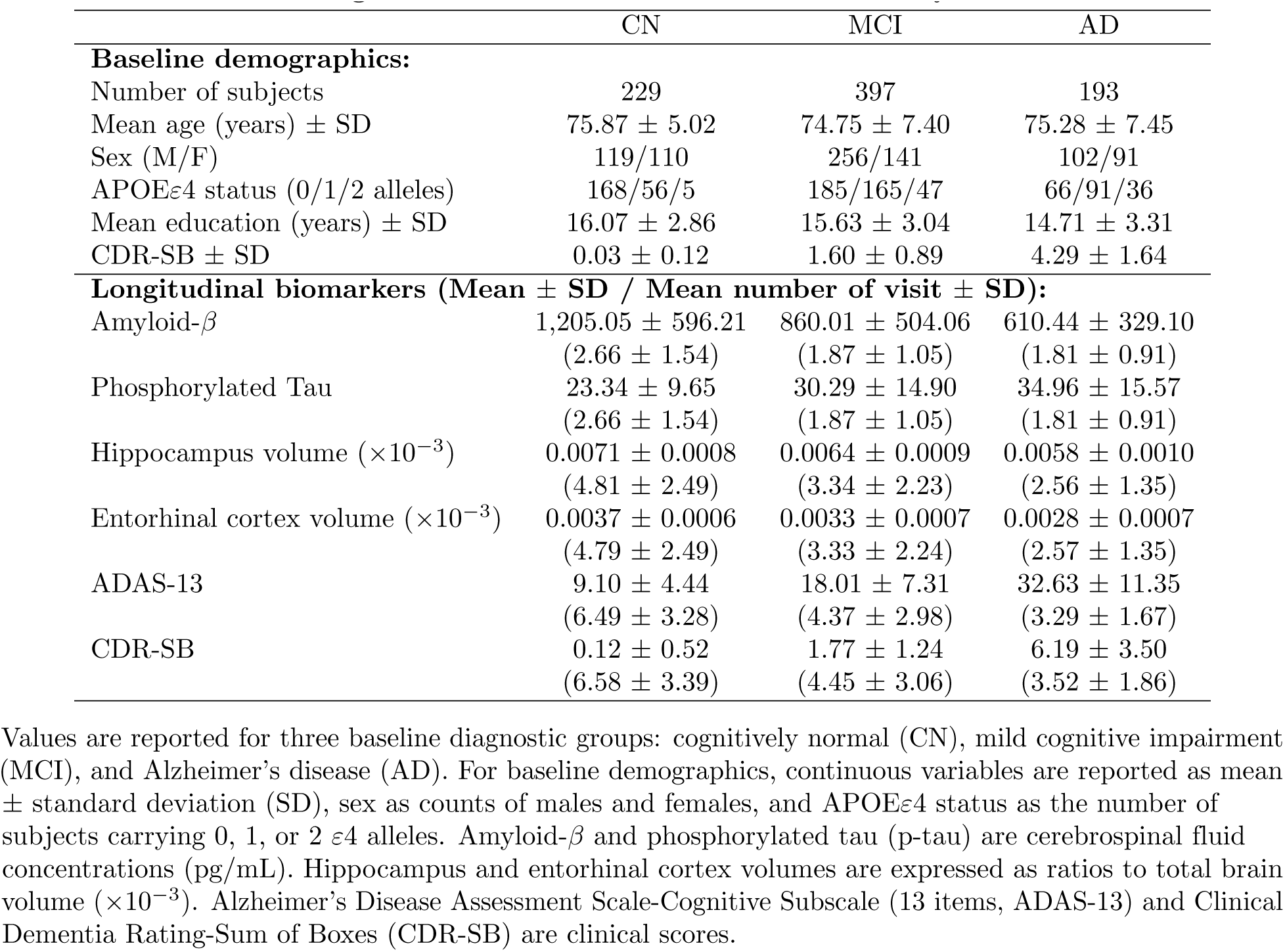
Baseline and longitudinal characteristics of the ADNI study cohort.

|  | CN | MCI | AD |
| --- | --- | --- | --- |
| **Baseline demographics:** |  |  |  |
| Number of subjects | 229 | 397 | 193 |
| Mean age (years) ± SD | 75.87 ± 5.02 | 74.75 ± 7.40 | 75.28 ± 7.45 |
| Sex (M/F) | 119/110 | 256/141 | 102/91 |
| APOEε4 status (0/1/2 alleles) | 168/56/5 | 185/165/47 | 66/91/36 |
| Mean education (years) ± SD | 16.07 ± 2.86 | 15.63 ± 3.04 | 14.71 ± 3.31 |
| CDR-SB ± SD | 0.03 ± 0.12 | 1.60 ± 0.89 | 4.29 ± 1.64 |
| **Longitudinal biomarkers (Mean ± SD / Mean number of visit ± SD):** |  |  |  |
| Amyloid-β | 1,205.05 ± 596.21<br>(2.66 ± 1.54) | 860.01 ± 504.06<br>(1.87 ± 1.05) | 610.44 ± 329.10<br>(1.81 ± 0.91) |
| Phosphorylated Tau | 23.34 ± 9.65<br>(2.66 ± 1.54) | 30.29 ± 14.90<br>(1.87 ± 1.05) | 34.96 ± 15.57<br>(1.81 ± 0.91) |
| Hippocampus volume ($\times 10^{-3}$) | 0.0071 ± 0.0008<br>(4.81 ± 2.49) | 0.0064 ± 0.0009<br>(3.34 ± 2.23) | 0.0058 ± 0.0010<br>(2.56 ± 1.35) |
| Entorhinal cortex volume ($\times 10^{-3}$) | 0.0037 ± 0.0006<br>(4.79 ± 2.49) | 0.0033 ± 0.0007<br>(3.33 ± 2.24) | 0.0028 ± 0.0007<br>(2.57 ± 1.35) |
| ADAS-13 | 9.10 ± 4.44<br>(6.49 ± 3.28) | 18.01 ± 7.31<br>(4.37 ± 2.98) | 32.63 ± 11.35<br>(3.29 ± 1.67) |
| CDR-SB | 0.12 ± 0.52<br>(6.58 ± 3.39) | 1.77 ± 1.24<br>(4.45 ± 3.06) | 6.19 ± 3.50<br>(3.52 ± 1.86) |

Values are reported for three baseline diagnostic groups: cognitively normal (CN), mild cognitive impairment (MCI), and Alzheimer’s disease (AD). For baseline demographics, continuous variables are reported as mean ± standard deviation (SD), sex as counts of males and females, and APOE $\epsilon$ 4 status as the number of subjects carrying 0, 1, or 2 $\epsilon$ 4 alleles. Amyloid- $\beta$ and phosphorylated tau (p-tau) are cerebrospinal fluid concentrations (pg/mL). Hippocampus and entorhinal cortex volumes are expressed as ratios to total brain volume ( $\times 10^{-3}$ ). Alzheimer’s Disease Assessment Scale-Cognitive Subscale (13 items, ADAS-13) and Clinical Dementia Rating-Sum of Boxes (CDR-SB) are clinical scores.

**Table 2.** Number of observed measurements per biomarker in each data split.

| Biomarker | Training | Test | Validation |
| --- | --- | --- | --- |
| Amyloid-β | 921 | 68 | 89 |
| Phosphorylated Tau | 921 | 68 | 89 |
| Hippocampus | 2,804 | 296 | 307 |
| Entorhinal cortex | 2,790 | 289 | 307 |
| ADAS-13 | 3,935 | 529 | 594 |
| CDR-SB | 4,022 | 596 | 610 |

Measurement counts for the training, test, and validation sets. The test set comprises the held-out final visit of each training subject (temporal hold-out), whereas the validation set comprises entirely unseen subjects (subject-level hold-out).

#### Data preparation

Data used in this study are preprocessed as follows:

##### Age

Age at visit in ADNI-1 ranged from 54 to 104 years (mean 78.32 years). For the Disease Progression Score (DPS; see the Disease Progression Score (DPS) section), age *t* was standardised to a reference of 75 years with a scale of 10 years (Eq 1), placing most values within approximately [*−*2.5, 2.5]. Centring on 75 years rather than the empirical mean keeps the DPS interpretable relative to the reference age used for the onset parameter (*β*). A wide scale reduces the sensitivity of the progression rate parameter (*α*), yielding smoother biomarker dynamics, more stable optimisation, and avoiding unrealistically steep trajectories.

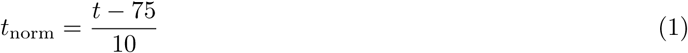

##### Biomarkers

Each biomarker was scaled to [0, 1] by min-max normalisation, with the minimum and maximum captured across the study population. For biomarkers that decrease with progression (amyloid-*β*, hippocampus volume, and entorhinal cortex volume), the normalised values were inverted so that all trajectories increase monotonically in the cascade model (Eq 2).

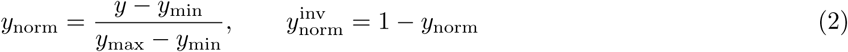

##### Baseline demographics

Sex was encoded as a binary (0 = male, 1 = female). APOE*ε*4 status was encoded as 0, 1, or 2, corresponding to non-carriers, heterozygotes (one *ε*4 allele), and homozygotes (two *ε*4 alleles), respectively. Baseline age was standardised as in Eq 1. Years of education and baseline CDR-SB were min-max normalised.

#### Disease Progression Score (DPS)

The latent disease timeline is defined using the Disease Progression Score (DPS) framework [6]. Each subject is assigned a progression rate (*α_i_*) and an onset time (*β_i_*) that together time-warp chronological age, aligning that individual’s longitudinal observations along a common latent disease scale. For subject *i* at chronological age *t*, the DPS *s_i_*(*t*) is a linear function of standardised age:

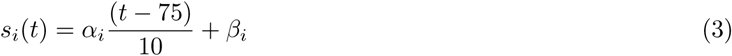

The progression rate *α_i_* is strictly positive (*α_i_ >* 0), enforced by an exponential parameterisation (see the Variational extension section). As a reference point, *α_i_* = 1 corresponds to a one-unit increase in DPS per decade of chronological time, with larger values indicating faster progression. The onset time *β_i_* is the subject’s DPS at the reference age of 75 years; *β >* 0 indicates earlier disease onset relative to the cohort average, and *β <* 0 indicates later onset.

#### Variational extension

To capture inter-individual variability and quantify predictive uncertainty, we extend the DPS framework by treating the subject-specific parameters *α_i_* and *β_i_* as probabilistic rather than point estimates. We assume that a subject’s progression rate and onset can be partially predicted from baseline demographics and clinical features (**X***_i_*). In this work, we restrict **X***_i_* to include only simple, non-invasive observation variables, including age, sex, year of education, APOE*ε*4 status, and baseline CDR-SB. To guarantee a positive progression rate, the rate is modelled in log-space. The encoder outputs the parameters of a Gaussian approximate posterior over log *α_i_*, and *α_i_* is recovered by exponentiation. The onset time *β_i_*is modelled directly as Gaussian. For each subject:

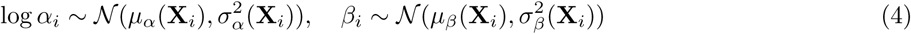

so that *α_i_ >* 0 and *α_i_* follows a Log-normal distribution. These posterior distributions are learned by variational inference [28, 29]. The encoder maps **X***_i_* to the distributional parameters 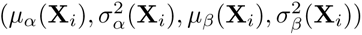 for each subject (Fig 1)

To allow gradients to propagate through the sampling step during training, the parameters are drawn using the reparameterisation trick [29]:

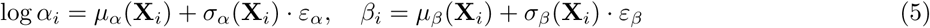

where *α_i_* = exp(log *α_i_*) and *ε_α_, ε_β_ ∼ N* (0, 1). Given the sampled parameters (*α_i_, β_i_*), the disease progression score is then computed using Eq 3 and subsequently passed to the biomarker trajectory model. Propagating samples in this way carries the uncertainty in *α_i_* and *β_i_* forward into the predicted biomarker trajectories, enabling uncertainty quantification for each individual’s disease trajectory (see the Uncertainty analysis section).

### Biomarker trajectory model

The population-level biomarker trajectories are structured according to the amyloid-*β* cascade hypothesis [9, 30, 31] (Fig 2). Under this hypothesis, amyloid-*β* (*A_β_*) accumulation precedes and drives phosphorylated tau pathology (*τ_p_*), which in turn drives neurodegeneration in the hippocampus (*N*_1_) and entorhinal cortex (*N*_2_). These changes ultimately lead to cognitive decline, as reflected in ADAS-13 (*C*_1_) and CDR-SB (*C*_2_). We encode this ordering as a fixed cascade topology and estimate the strength of each link from data.

**Fig 2.**
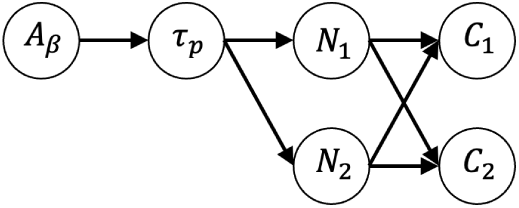
Cascade structure of the biomarker trajectory model. Amyloid-*β* (*A_β_*) accumulation drives phosphorylated tau pathology (*τ_p_*), which drives neurodegeneration in the hippocampus (*N*_1_) and entorhinal cortex (*N*_2_), both of which in turn drive cognitive decline as measured by ADAS-13 (*C*_1_) and CDR-SB (*C*_2_) scores. The strength of each edge (*λ*) is estimated from the data, while the topology itself is fixed.

Because AD progression typically follows a pattern of acceleration followed by a plateau, each biomarker trajectory *y_k_*(*s*) is modelled as a logistic function of DPS. This captures the direct effect of DPS on the biomarker *k*, together with the influence of its parent biomarkers in the cascade. The full cascade model can thus be expressed as:

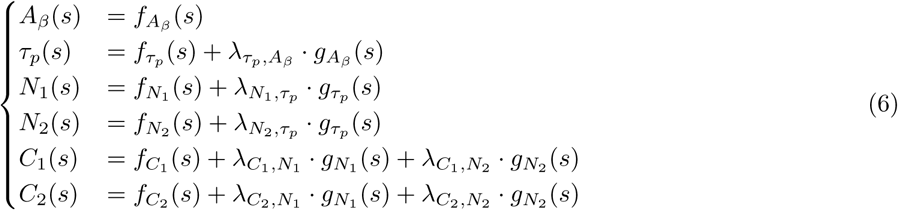

where the direct effect is the logistic function

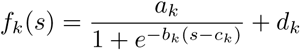

in which *c_k_*is the DPS at which biomarker *k* reaches the midpoint of its pathological change, *b_k_*controls steepness, *a_k_* the dynamic range, and *d_k_* a linear offset. The contribution of a parent biomarker is modelled as

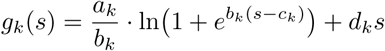

which is the integral of *f_k_*(*s*). Using this integral means that a child biomarker responds to the accumulated burden of its parent up to disease stage *s*, rather than the parent’s instantaneous level, reflecting the cumulative nature of neuropathological damage. The parameter *λ* quantifies cascade strength. Importantly, the parameters (*a_k_, b_k_, c_k_, d_k_*) are shared between *f_k_*(*s*) and *g_k_*(*s*), ensuring consistency between the direct logistic trajectory of biomarker *k* and the cumulative effect it exerts on downstream biomarkers.

### Training procedure

The framework is trained by alternating two optimisation steps, a variational analogue of the Expectation-Maximisation (EM) algorithm [32], until convergence. This jointly optimises the subject-specific latent parameters (*α_i_, β_i_*) and the population-level cascade parameters (*θ_k_*) while propagating individual uncertainty. In the E-step, the encoder is updated to improve the subject-specific posterior given the current cascade model. In the M-step, the cascade parameters are updated given the current posteriors over (*α_i_, β_i_*). Because inference is amortised through the encoder, which maps baseline features directly to posteriors, the trained model can produce individualised posteriors for previously unseen subjects without re-optimisation.

The framework was implemented in PyTorch and optimised with Adam, using learning rates of 0.005 (E-step) and 0.01 (M-step), with adaptive learning rate schedulers. The model was trained with a batch size of 100 for up to 500 epochs, with early stopping.

#### Encoder update (E-step) — inferring subject-specific parameters

In the E-step, the cascade parameters *θ_k_* are held fixed while the encoder is updated. The encoder maps the 5 baseline features to a 32-dimensional hidden representation via a 3-layer fully connected network (with ReLU activations, batch normalisation, and dropout), and outputs the mean and standard deviation of the approximate posteriors over log *α_i_* and *β_i_*.

The encoder is trained by minimising the negative Evidence Lower Bound (ELBO) [29]. The objective function (*L_E_*) comprises a reconstruction term, given by the negative log-likelihood (NLL), and a regularisation term, given by the Kullback-Leibler (KL) divergence [33] between the variational posterior *q_i_* = *q*(*α_i_, β_i_ |* **X***_i_*) and the predefined prior *p*. For subject *i*, visit *j* and biomarker *k*, the loss is defined as:

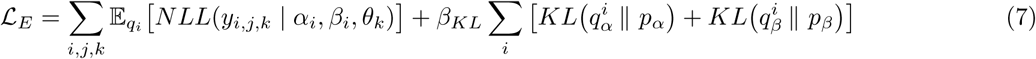

where 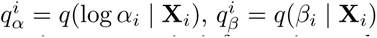 and *β_KL_* = 0.1 weights the KL term. This down-weighting allows the posteriors to remain informative and subject-specific rather than collapsing toward the prior. The prior over log *α_i_* is Gaussian, equivalently, a log-normal prior (*LN* (1.2, 0.5^2^)). Positivity of *α_i_* is guaranteed by the exponential parametrisation (see Variational extension section). The prior over *β_i_* is Gaussian *N* (0.0, 8.0^2^), permitting a wide range of onset times.

#### Cascade model update (M-step) — fitting population-level trajectories

In the M-step, the encoder is held fixed and the population-level parameters, the cascade parameters *θ_k_* = *{a_k_, b_k_, c_k_, d_k_, λ_k,parent_}* and the modality-specific aleatoric noise variance 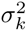, are updated using the current posteriors over (*α_i_, β_i_*). The objective function is:

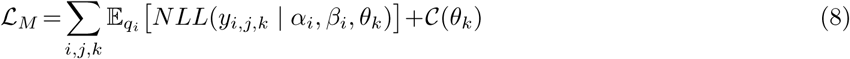

where *C*(*θ_k_*) penalises trajectories that leave clinically plausible, normalised bounds. Predicted values at the extremes of the DPS axis are constrained so that *ė_k_*(*−*10) *∈* [0, 0.45] (near-normal at the earliest stage) and *ė_k_*(10) *∈* [0.55, 1.0] (near-saturated at the most advanced stage):

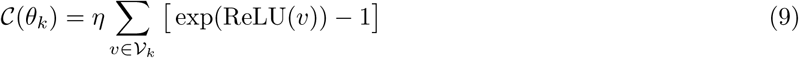

with 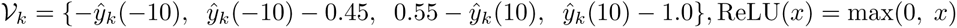, and regularisation weight *η* = 2.0. The penalty is zero when predictions lie within the bounds and grows exponentially with the degree of violation.

To keep trajectories monotonically increasing across disease progression, the logistic growth rate *b_k_*is constrained to be positive. Following [6], whenever *b_k_ <* 0, the parameters are reparameterised as *a_k_* = *−a_k_*, *b_k_* = *−b_k_*, and *d_k_* = *d_k_* + *a_k_*. This transformation leaves *f_k_*(*s*) unchanged while ensuring *b_k_ >* 0, so monotonicity is imposed without altering the fitted trajectory.

#### Sparse data handling

The ADNI dataset is highly sparse, as not every biomarker is measured at each visit. In our training set, for example, CSF biomarkers (amyloid-*β* and p-tau) were available at only 921 of 4,055 visits, as shown in Table 2. During model training, the loss functions defined for the E-step (*L_E_*, Eq 7), M-step (*L_M_*, Eq 8), and personalised fine-tuning (*L_P_*, Eq 12) are computed only over the available biomarkers at each visit. This approach ensures that unobserved biomarkers do not influence the optimisation, while allowing the model to utilise all available information.

### Uncertainty analysis

Baseline features (**X***_i_*) are encoded into a variational posterior distribution over the latent parameters (*α_i_, β_i_*), capturing subject-level uncertainty. This uncertainty is propagated through the DPS and into the biomarker trajectories:

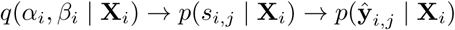

The total predictive uncertainty comprises two distinct components: epistemic uncertainty and aleatoric uncertainty [34]. Epistemic (model) uncertainty reflects incomplete knowledge of the true subject-specific parameters. It is estimated by drawing Monte Carlo (MC) samples of (*α_i_, β_i_*) from the variational posterior, propagating each through the cascade, and taking the standard deviation of the resulting biomarker predictions (*σ_epi_*). It can be reduced as new observations become available. Aleatoric (data) uncertainty represents the inherent measurement noise in clinical assessments and is captured by a modality-specific observation-noise term, optimised during the M-step (see Cascade model update (M-step) — fitting population-level trajectories section).

For the fluid (amyloid-*β* and p-tau) and structural neuroimaging (hippocampus and entorhinal cortex) biomarkers, the aleatoric observation noise is modelled as homoscedastic, remaining constant across the disease course. For these modalities, its standard deviation is initialised at 0.1 and optimised in the M-step.

Cognitive assessments are measured on bounded scales (for example, the CDR-SB ranges from 0 to 18, with higher scores indicating greater impairment), and their measurement range tends to widen as impairment progresses. We therefore model the aleatoric noise for the cognitive measures (ADAS-13 and CDR-SB) as heteroscedastic, with a standard deviation that varies log-linearly with the DPS:

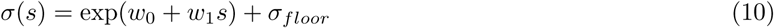

Each cognitive biomarker has its own parameters (*w*_0_*, w*_1_*, σ_floor_*), initialised to -3.0, 0.1, and 0.1, respectively, and optimised jointly with the population-level trajectory parameters in the M-step. The positive initial *w*_1_ encodes increasing noise with progression, while *σ_floor_* imposes a lower bound on the observation noise.

Assuming independence between the epistemic and aleatoric components, the total predictive standard deviation for each biomarker is:

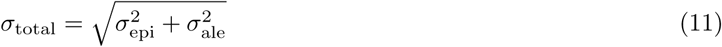

The framework thus provides, for each subject and visit, a point estimate together with a credible interval (CrI), enabling prognosis for previously unseen subjects from baseline measurements alone.

### Personalised fine-tuning

Predicted trajectories can be progressively refined as longitudinal observations become available, using an approximate sequential Bayesian updating scheme in which the posterior inferred from earlier visits serves as the prior for the current visit. For each subject, the subject-specific posterior over (*α_i_, β_i_*) is fine-tuned on the available longitudinal data by minimising at visit *j*.

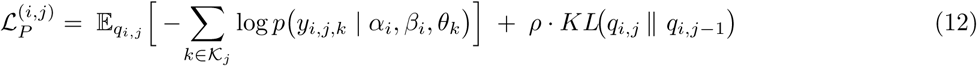

where *q_i,j_* = *q*(*α_i_, β_i_ |* **y***_i,_*_1:*j*_, **X***_i_*) is the posterior distribution updated using all visits up to *j*, and *q_i,j−_*_1_ = *q*(*α_i_, β_i_ |* **y***_i,_*_1:*j*_*_−_*_1_, **X***_i_*) is the posterior from the preceding visits. *K_j_* is the set of biomarkers observed at visit *j* and the population-level cascade parameters *θ_k_*are held fixed at their trained values. The first term is the reconstruction loss over the biomarkers observed at visit *j*.The second term anchors the updated posterior to the previous one, which, at the first update, is the baseline population-informed posterior *q*(*α_i_, β_i_ |* **X***_i_*). The weight *ρ* controls this anchoring and is annealed with the number of observations:

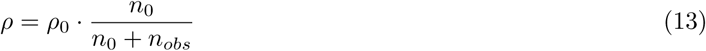

where *ρ*_0_ = 0.1 is the initial weight, *n*_0_ is a pseudo-count encoding the strength of the population prior, and *n_obs_* is the number of measurements observed for the subject. When *n_obs_*is small, *ρ* is large, and the estimate stays close to the population-informed posterior, guarding against overfitting to sparse early data. As observations accumulate, *ρ* shrinks, and the estimate is personalised.

During fine-tuning, the encoder’s hidden layers are frozen, and only the four output heads corresponding to the mean and standard deviations of the posteriors over log *α_i_* and *β_i_* are updated. Fine-tuning runs for up to 50 epochs, with early stopping after 10 consecutive epochs without improvement. This sequential updating allows the model to adapt as new data arrive, reflecting the real-world clinical process in which clinicians progressively refine diagnoses and prognoses in light of new longitudinal findings.

## Results

We evaluated the framework on longitudinal ADNI data along three axes: the stability and biological plausibility of the inferred disease timeline, the recovered cascade strengths and biomarker ordering, and the accuracy and calibration of individualised uncertainty-aware predictions for previously unseen subjects. Unless stated otherwise, results are reported across 20 independent training runs with different random seeds.

### Training losses

The framework was trained using the two-step algorithm described in the Training procedure section, jointly learning the population-level trajectories and the subject-specific parameters (*α_i_, β_i_*). To assess optimisation stability, we repeated training 20 times with different random seeds. All runs converged to comparable optima. The best total loss was *−*1.253 *±* 0.051 (mean *±* standard deviation), with the reconstruction, cascade, and KL components showing low variability across runs, indicating robustness to initialisation (see S1 Fig and S1 Table).

### Population-level biomarker trajectories

The model aligned heterogeneous, asynchronous observations onto a common disease timeline defined by the DPS, yielding smooth, monotonic biomarker trajectories consistent with the known course of Alzheimer’s disease (Fig 3). Notably, although the clinical diagnoses (cognitively normal, CN; mild cognitive impairment, MCI; and Alzheimer’s disease, AD) were not provided as model inputs, the inferred DPS separated the three groups in the expected order (Fig 4a). This demonstrates an unsupervised alignment with clinical staging that the model was never explicitly trained to reproduce.

**Fig 3.**
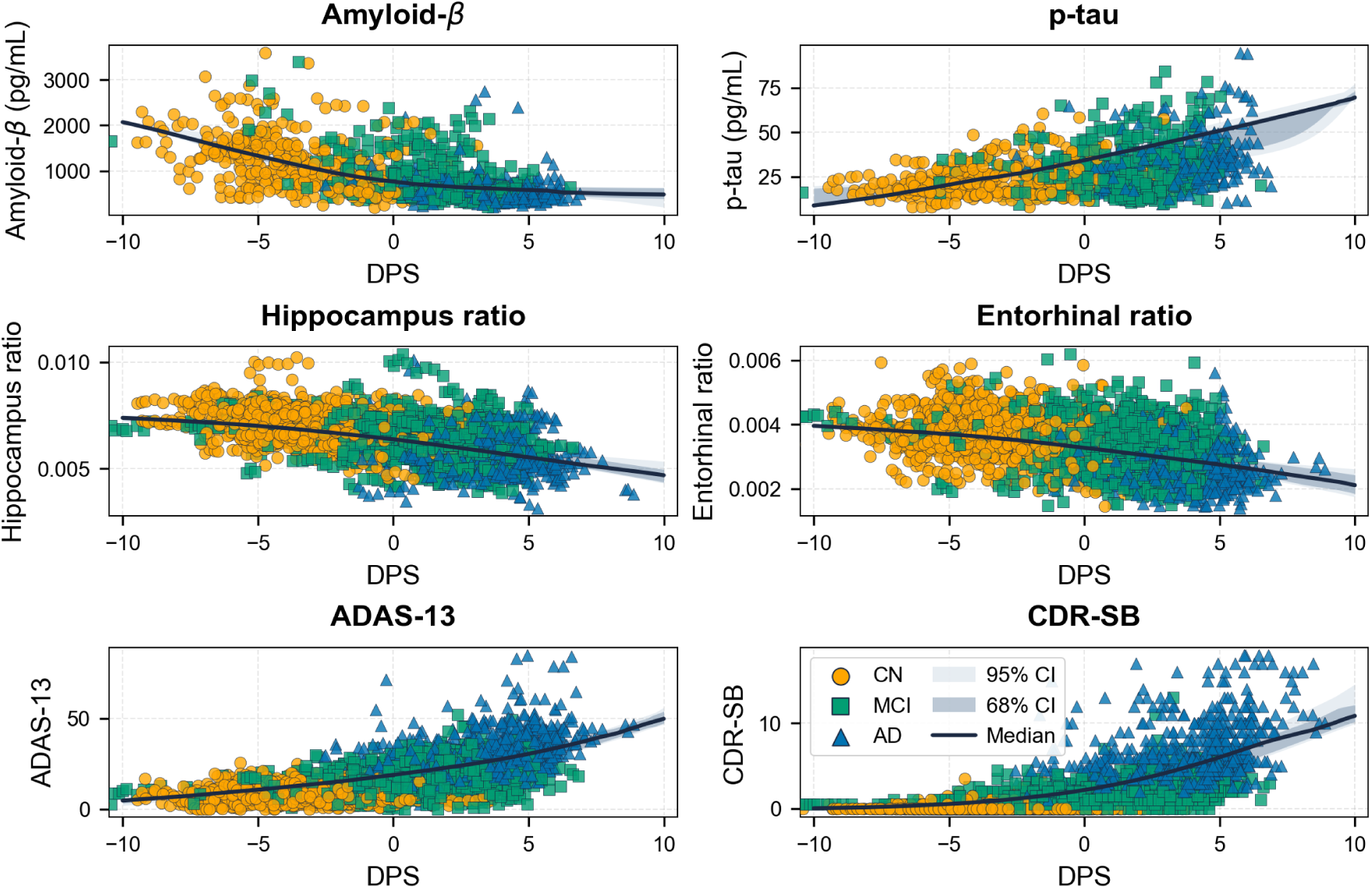
Population-level biomarker trajectories along the inferred disease timeline. Biomarker trajectories estimated by the cascade functions of the Disease Progression Score (DPS) using the training dataset. Each point represents an individual observation, coloured by the clinical diagnosis at the corresponding visit (cognitively normal, CN; mild cognitive impairment, MCI; and Alzheimer’s disease, AD). The solid black curve shows the mean fitted trajectory across 20 independent training runs, and the shaded region indicates the corresponding 68% and 95% confidence intervals.

**Fig 4.**
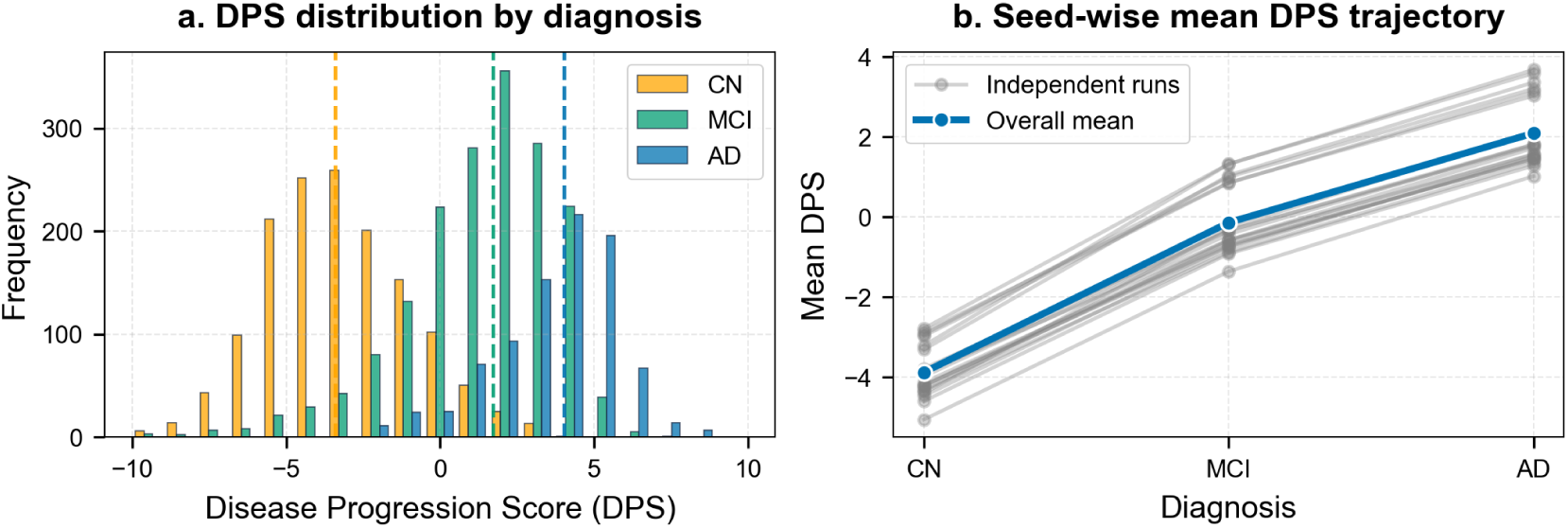
Distribution and stability of the latent Disease Progression Score (DPS) across diagnostic groups. **(a)** Histogram showing the distribution of DPS values stratified by clinical diagnosis (CN, MCI, AD) from a representative training run. Substantial overlap, particularly for MCI, reflects the continuous biological spectrum of Alzheimer’s disease progression. **(b)** Stability of group-level DPS estimates across 20 independent training runs. Grey markers represent the mean DPS for each diagnostic group in each run, while blue markers denote the overall mean across runs, demonstrating consistent separation between diagnostic groups despite different random initialisations.

The DPS distribution across diagnostic groups showed a progressive shift with increasing severity (Fig 4a). We further analyse the separability between diagnostic groups using the pairwise area under the ROC curve (AUC), which measures the probability that a more severe diagnosis is assigned a higher DPS than a less severe one. The AUC computed within each run and averaged across 20 seeds was 0.98 *±* 0.003 for CN versus AD, 0.898 *±* 0.009 for CN versus MCI, and 0.785 *±* 0.006 for MCI versus AD (mean*±*SD). The strong CN-AD separation confirms that the inferred timeline recovers the clinical ordering, whereas the lower MCI-AD value quantifies the overlap visible in Fig 4a, with MCI spanning a broad DPS range that bridges the CN and AD distributions. The small across-seed variation in these values indicates that the separation is itself stable to initialisation. This pattern is consistent with the recognised biological heterogeneity of the MCI stage [35] and the continuous nature of Alzheimer’s disease progression.

To assess stability, we repeated the fit across 20 independent training runs (Fig 4b). The group ordering was preserved in every run: the group-mean DPS was -3.88 *±* 0.64 for CN, -0.15 *±* 0.83 for MCI, and 2.09 *±* 0.85 for AD (mean*±*SD). This consistent separation indicates that the inferred DPS is a stable, continuous staging measure that recovers the clinical ordering while supporting longitudinal tracking and cross-subject comparison.

Along the inferred timeline, biomarkers reached their pathological midpoints, the DPS at which each attains 50% of its modelled change, in an order broadly consistent with established Alzheimer’s disease staging (Fig 5). Amyloid-*β* reached its midpoint earliest and consistently the earliest across seeds (mean *±* SD: *s* = *−*4.77 *±* 1.65), followed by the medial temporal and cognitive measures. Notably, while the cascade topology is defined a priori, it acts as a soft constraint: it encourages but does not strictly enforce this broad sequential progression across biomarker modalities. The p-tau midpoint was the least stable across seeds (*s* = 1.36 *±* 2.81), likely reflecting the protracted dynamics of tau pathology, which accumulates gradually over an extended period, estimated at roughly 15–20 years to reach typical AD levels [36]. Hence, the pathology midpoint is intrinsically difficult to localise along the latent timeline. Its position relative to the neurodegeneration markers should therefore be interpreted with caution.

**Fig 5.**
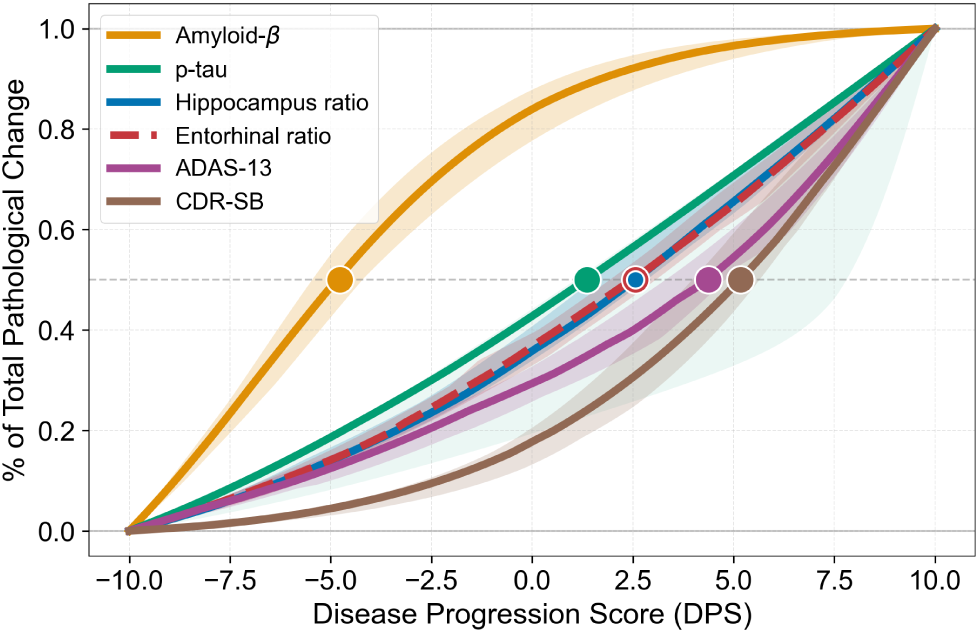
Temporal ordering of Alzheimer’s disease biomarkers along the inferred disease timeline. The mean proportion of pathological change for each biomarker is shown as a function of Disease Progression Score (DPS). The curve represents the average across 20 independent training runs, and the shaded bands represent the *±*1 standard deviation across runs. The dashed horizontal line indicates the 50% level of maximum change (the pathological midpoint), consistent with established staging. Note that the hippocampus and entorhinal midpoints coincide (both at *s* = 2.56) and are shown as a single overlaid marker.

The more informative, data-driven results are the relative timing of biomarkers at the same cascade level, where the topology does not dictate their sequence. Hippocampal and entorhinal atrophy, parallel downstream targets of tau, reached their midpoints synchronously (*s* = 2.56 *±* 0.66 and *s* = 2.56 *±* 0.69; mean per-seed difference 0.10 DPS units), matching the spatiotemporal pattern of medial temporal lobe neurodegeneration [37, 38]. Among the cognitive measures, ADAS-13 tended to reach its midpoint before CDR-SB (*s* = 4.37 *±* 0.89 and *s* = 5.18 *±* 0.71; ADAS-13 earlier in 16 of 20 seeds), consistent with cognitive testing detecting impairment earlier than global functional staging [39].

### Subject-specific parameters and feature associations

To characterise inter-individual heterogeneity in disease progression, we examined associations between baseline clinical features and the model-derived subject-specific parameters: progression rate (*α*) and onset time (*β*). As noted before, *α* controls the rate of disease progression along the Disease Progression Score (DPS), with a one-unit increase in *α* corresponding to a one-unit increase in the DPS every 10 years. *β* determines the relative timing of disease onset. Higher *β* values correspond to earlier inferred onset time, whereas negative values indicate later-onset trajectories relative to the baseline cognitive age of 75 years. Notably, the associations outlined in this section specifically describe the mapping learned by the encoder.

APOE*ε*4 was the strongest determinant of the inferred trajectories (Fig 6, Table 3, 4). Carriers progressed faster than non-carriers, and increasing allele count was associated with markedly earlier onset. APOE*ε*4 genotype alone explained roughly a third of the variance in inferred onset (*η*^2^ = 0.35), with mean *β* rising from *−*3.17 in non-carriers to +3.16 in homozygotes (Table 4). These findings are consistent with extensive clinical evidence demonstrating that APOE*ε*4 accelerates pathological accumulation and shifts the age of onset earlier in Alzheimer’s disease [40].

**Fig 6.**
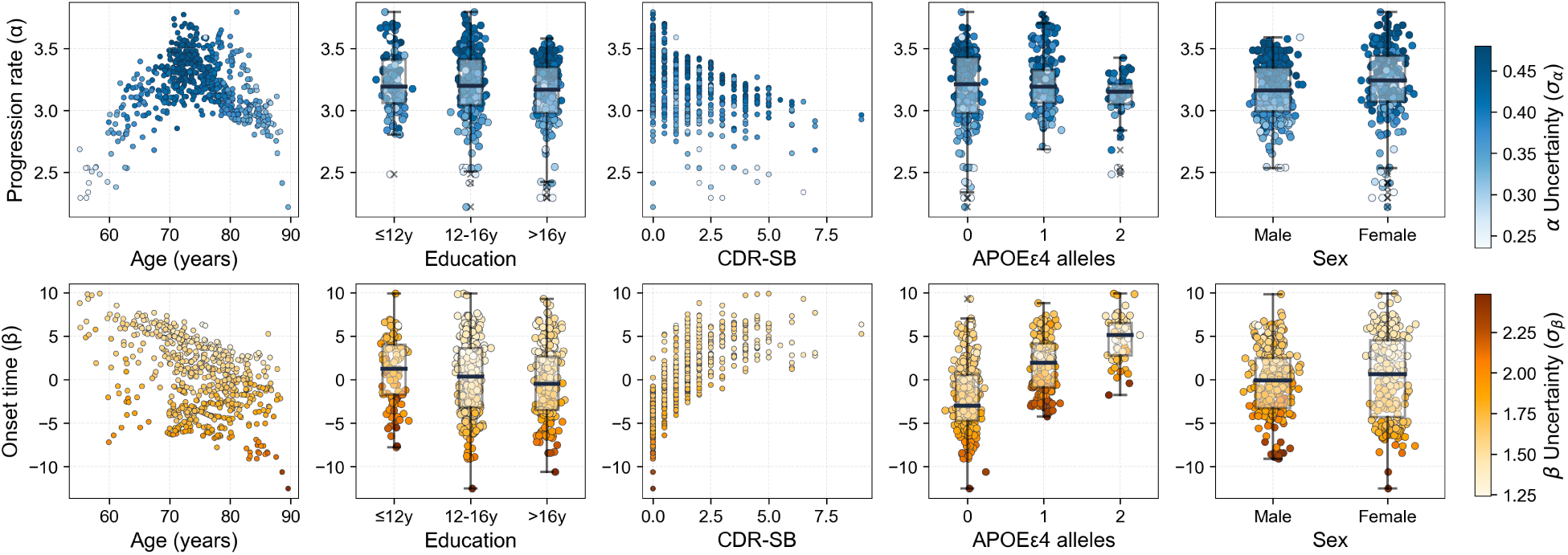
Associations of baseline features with subject-specific parameters. The inferred progression rate (*α*, top row) and onset time (*β*, bottom row) are evaluated against baseline demographic and clinical features. Scatter plots show the relationships for continuous features (age and CDR-SB). Box plots present group-level comparisons for categorical features (education, APOE*ε*4 status and sex). Across all panels, colour intensity of data points denotes the model-derived posterior uncertainty (*σ*) for each individual’s parameter estimate. Data are visualised from a representative run.

**Table 3.** Association between baseline features and subject-specific progression parameters.

| Feature | Progression rate<br>($\mu_\alpha$) | Onset time<br>($\mu_\beta$) | Uncertainty<br>Progression rate ($\sigma_\alpha$) | Uncertainty<br>Onset time($\sigma_\beta$) |
| --- | --- | --- | --- | --- |
| *Continuous features – Pearson $r$ (mean ± SD)* |  |  |  |  |
| Age | $-0.083 \pm 0.082$ | **$-0.494 \pm 0.025$** | $-0.173 \pm 0.057$ | $0.251 \pm 0.248$ |
| CDR-SB | $-0.188 \pm 0.161$ | **$0.727 \pm 0.010$** | $-0.088 \pm 0.088$ | $-0.132 \pm 0.394$ |
| *Categorical features – ANOVA $\eta^2$ (mean ± SD); (seeds sig., $p < 0.05$)* |  |  |  |  |
| Education | $0.022 \pm 0.018$<br>(15/20) | $0.013 \pm 0.002$<br>(19/20) | $0.005 \pm 0.005$<br>(5/20) | $0.031 \pm 0.017$<br>(18/20) |
| APOEε4 | **$0.079 \pm 0.050$**<br>(20/20) | **$0.345 \pm 0.014$**<br>(20/20) | $0.025 \pm 0.014$<br>(18/20) | $0.128 \pm 0.117$<br>(18/20) |
| *Binary categorical features – two-sample $t$-test; (seeds sig., $p < 0.05$)* |  |  |  |  |
| Sex | (14/20) | (8/20) | (14/20) | (10/20) |

Association between baseline features and the inferred progression rate ( $\mu_\alpha$ ), onset time ( $\mu_\beta$ ), and their respective uncertainties ( $\sigma_\alpha, \sigma_\beta$ ) across 20 independent training runs. Continuous features (age and CDR-SB) are evaluated using the Pearson correlation coefficients ( $r$ , mean ± SD). Categorical features (education and APOE $\epsilon$ 4) are summarised using one-way ANOVA effect size ( $\eta^2$ , mean ± SD) and sex by a two-sample $t$ -test. For the categorical variables, the number of runs reaching significance at $p < 0.05$ is shown in parentheses. Bold denotes $|r| > 0.3$ (continuous) or $\eta^2 > 0.06$ with significance in all 20 runs (categorical).

**Table 4.** Subject-specific progression parameters for categorical variables.

| Baseline Feature | Progression rate<br>($\mu_\alpha$) | Onset time<br>($\mu_\beta$) | Uncertainty<br>Progression rate ($\sigma_\alpha$) | Uncertainty<br>Onset time($\sigma_\beta$) |
| --- | --- | --- | --- | --- |
| Education ≤ 12y | $3.159 \pm 0.055$ | $-0.302 \pm 0.806$ | $0.401 \pm 0.008$ | $1.522 \pm 0.080$ |
| Education 12 – 16y | $3.118 \pm 0.045$ | $-1.055 \pm 0.769$ | $0.399 \pm 0.006$ | $1.550 \pm 0.085$ |
| Education ≥ 16y | $3.068 \pm 0.039$ | $-1.572 \pm 0.731$ | $0.394 \pm 0.006$ | $1.577 \pm 0.095$ |
| APOEε4=0 | $3.051 \pm 0.059$ | $-3.172 \pm 0.692$ | $0.391 \pm 0.007$ | $1.580 \pm 0.118$ |
| APOEε4=1 | $3.169 \pm 0.039$ | $0.308 \pm 0.823$ | $0.405 \pm 0.006$ | $1.541 \pm 0.073$ |
| APOEε4=2 | $3.117 \pm 0.067$ | $3.160 \pm 0.885$ | $0.394 \pm 0.007$ | $1.513 \pm 0.062$ |
| Male | $3.079 \pm 0.041$ | $-1.478 \pm 0.768$ | $0.401 \pm 0.006$ | $1.558 \pm 0.097$ |
| Female | $3.133 \pm 0.048$ | $-0.933 \pm 0.742$ | $0.391 \pm 0.006$ | $1.560 \pm 0.080$ |

Values represent the group mean ± standard deviation across 20 independent training runs. Progression rate ( $\alpha$ ) and onset time ( $\beta$ ) capture individualised trajectory dynamics. Higher $\beta$ values indicate earlier onset.

Baseline CDR-SB was strongly associated with *β* (*r* = 0.73 *±* 0.01). As CDR-SB is itself a severity measure fed to the encoder, this largely reflects the model anchoring inferred onset to baseline clinical severity rather than an independent finding. Age was negatively associated with *β* (*r* = *−*0.494 *±* 0.03), indicating that older individuals tended to have later inferred onset. On the other hand, its relationship with *α* was near zero overall (*r* = *−*0.08 *±* 0.08) but appeared non-monotonic. In a representative run (Fig 6), *α* increased with age to approximately 70-75 years, reaching its peak in clinical aggressiveness. Then, the trend reverses into a negative correlation, with older individuals exhibiting slower and more variable progression rates. This pattern is consistent with clinical observations that mid-to-late 70s represent a peak window of rapid Alzheimer’s decline, whereas progression in the oldest population becomes increasingly heterogeneous, likely due to survivorship bias and the presence of fixed age-related pathologies [41].

Higher education was reproducibly but modestly associated with later onset (*β*). It was significant in 19 of 20 runs, though explaining only 1% of the variance in inferred onset (*η*^2^ = 0.01). This small, consistent effect aligns with the cognitive reserve hypothesis that education delays the emergence of clinical symptoms without altering underlying pathology [42]. Sex was not robustly associated with either parameter. Although some studies have reported faster progression in women, findings remain mixed and appear to be cohort-dependent [43].

Overall, posterior uncertainty showed no meaningful association with baseline features (Table 3). While statistically detectable differences in uncertainty for *α* and *β* were observed across APOE*ε*4 genotype, the effect size was negligible. Specifically, mean *σ_β_* decrease merely from 1.58 in non-carriers to 1.51 in homozygotes (Table 4). Thus, although genetic risk statistically contributes to variability in model confidence, its practical impact is minimal.

### Estimated cascade strengths

The model quantifies directed relationships between biomarkers through the cascade parameters *λ*, as detailed in Section Biomarker trajectory model. Because the cascade topology is fixed a priori, the directions of these links are imposed. The estimated strengths, however, are learned from data (Table 5). Among the seven links, the p-tau to neurodegeneration edges were estimated as the strongest (entorhinal 0.12 *±* 0.08, hippocampal 0.10 *±* 0.07). These findings indicate that tau pathology exerts the largest direct influence on medial temporal atrophy. This is consistent with the established link between tau burden and neuronal loss [44]. The amyloid-*β* to p-tau link was the weakest (0.025 *±* 0.007) yet the most reproducible of any edge (the lowest relative variability across runs), consistent with amyloid acting as a comparatively weak, slow upstream initiator of tau pathology rather than an immediate driver [45].

**Table 5.**
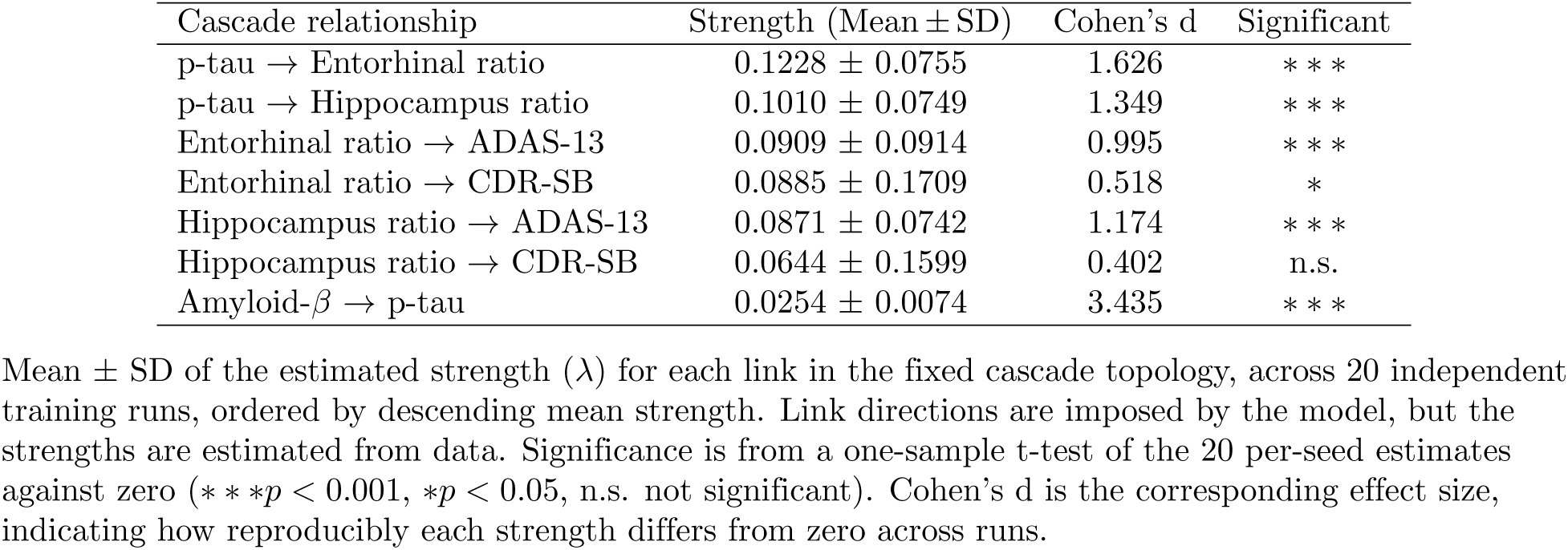
Estimated cascade strengths for the directed biomarker links.

| Cascade relationship | Strength (Mean ± SD) | Cohen’s d | Significant |
| --- | --- | --- | --- |
| p-tau → Entorhinal ratio | $0.1228 \pm 0.0755$ | 1.626 | *** |
| p-tau → Hippocampus ratio | $0.1010 \pm 0.0749$ | 1.349 | *** |
| Entorhinal ratio → ADAS-13 | $0.0909 \pm 0.0914$ | 0.995 | *** |
| Entorhinal ratio → CDR-SB | $0.0885 \pm 0.1709$ | 0.518 | * |
| Hippocampus ratio → ADAS-13 | $0.0871 \pm 0.0742$ | 1.174 | *** |
| Hippocampus ratio → CDR-SB | $0.0644 \pm 0.1599$ | 0.402 | n.s. |
| Amyloid-β → p-tau | $0.0254 \pm 0.0074$ | 3.435 | *** |

Mean ± SD of the estimated strength ( $\lambda$ ) for each link in the fixed cascade topology, across 20 independent training runs, ordered by descending mean strength. Link directions are imposed by the model, but the strengths are estimated from data. Significance is from a one-sample t-test of the 20 per-seed estimates against zero (\*\* $p < 0.001$ , \* $p < 0.05$ , n.s. not significant). Cohen’s d is the corresponding effect size, indicating how reproducibly each strength differs from zero across runs.

To assess which strengths were reliably estimated, we tested each edge against zero across the 20 runs using a one-sample t-test. Five of the seven links were reproducibly non-zero (*p <* 0.001). However, pathways linking atrophy of the hippocampus and entorhinal cortex to CDR-SB showed greater variability and weaker statistical support than their effects on the ADAS-13. This discrepancy likely reflects the distinct clinical domains captured by these assessments. Atrophy in the medial temporal lobe primarily affects episodic memory, which is more directly measured by ADAS-13, whereas CDR-SB captures broader functional abilities and daily living skills, leading to increased variability in the estimated cascade strength.

### Dynamic personalised prognosis with uncertainty

The proposed framework serves as a prognostic tool that generates individualised disease trajectories with explicitly quantified uncertainty. After population-level training, the model estimates subject-specific parameters—progression rate (*α*) and onset time (*β*)—for each individual using only their baseline clinical and demographic features. Importantly, the variational formulation naturally provides predictive uncertainty by sampling from the learned posterior distribution over these parameters, which is then propagated through the individual biomarker trajectory model (see Section Uncertainty analysis). The final optimised standard-deviation trajectories of aleatoric noise learned from the data are visualised in S3 Fig.

Moreover, the framework supports dynamic, subject-specific fine-tuning. As longitudinal clinical observations become available, the model updates the latent disease parameters to reflect the new data (see Section Personalised fine-tuning). As illustrated by a representative test subject (Fig 7), the inferred parameters were refined from their baseline estimates (*α* = 3.87, *β* = *−*3.11) to more precise posterior values (*α* = 5.31, *β* = 6.89). The updated estimates indicate a faster progression rate and markedly earlier onset than the baseline alone suggested. Consequently, parameter uncertainty was reduced: uncertainty regarding the progression rate decreased by 66.4% (*σ_α_* from 1.861 to 0.625), and uncertainty over the delayed onset decreased modestly by 6.5% (*σ_β_* from 1.71 to 1.60). The follow-up data primarily constrained the slope of decline rather than the absolute timing of onset. Correspondingly, the credible intervals for each biomarker narrowed substantially, except for the cognitive measures, reflecting their larger variability at the later stage of the disease.

**Fig 7.**
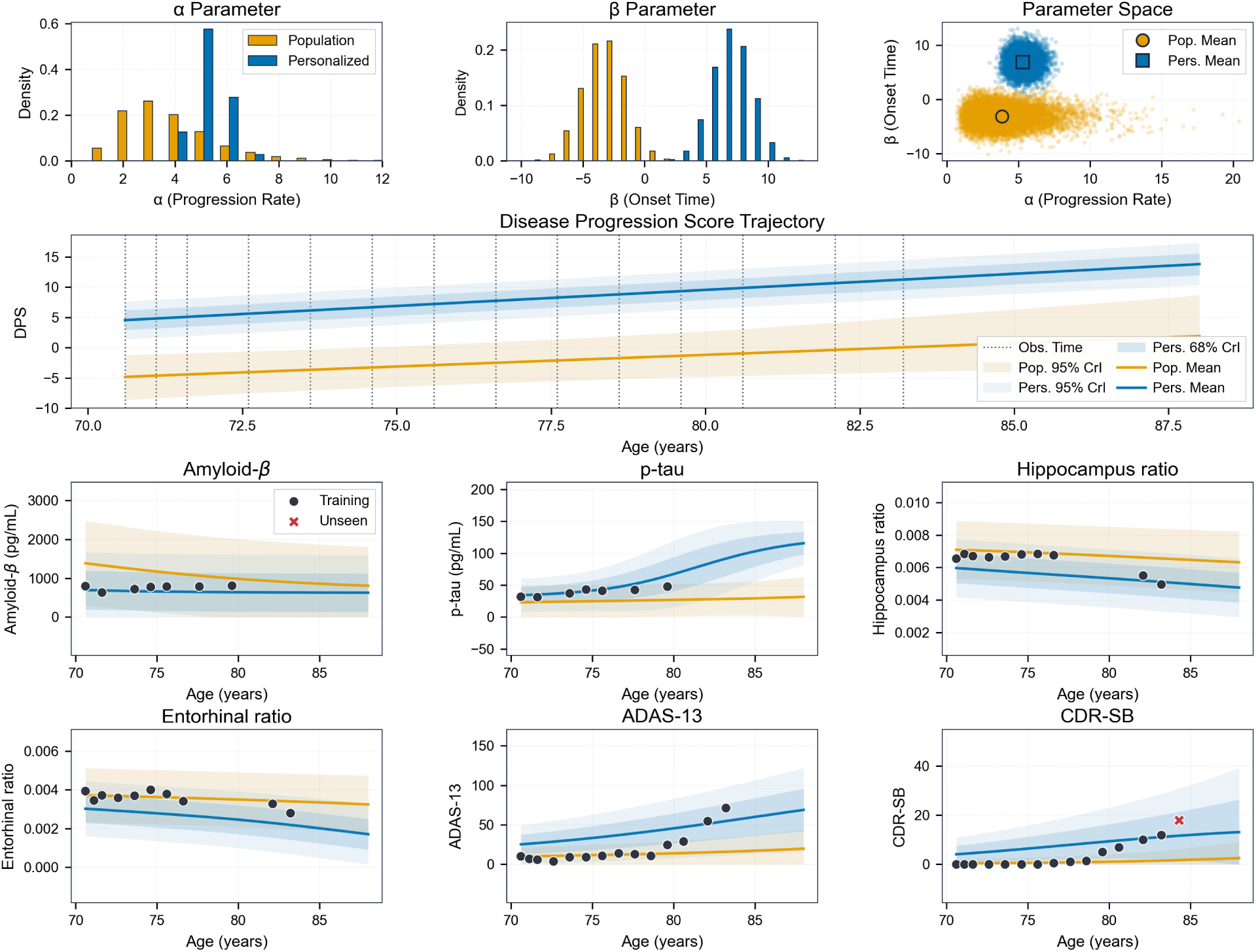
Dynamic fine-tuning refines individualised prognosis and reduces predictive uncertainty. A representative test subject demonstrates the framework’s capacity to update trajectory forecasts using individual longitudinal observations. Throughout all panels, orange represents prior estimates (derived from baseline features), and blue denotes the refined posterior (personalised) estimates after incorporating longitudinal data. **Top row**: Probability distributions of the latent progression rate (*α*, left) and onset time (*β*, middle), alongside their joint parameter space (right). **Middle row**: The corresponding Disease Progression Score (DPS) trajectory across chronological age. Vertical dotted lines indicate the chronological timing of the observed clinical visits. **Bottom rows**: Longitudinal biomarker trajectory predictions. Shaded regions represent 68% and 95% credible intervals (CrI). Black dots represent the observed clinical visits used for model fine-tuning, and the red crosses indicate unseen test visits.

At the group level, subject-specific fine-tuning improved point predictions for most biomarkers relative to the baseline population-level model (Table 6, S2 Fig), with the largest relative reduction in root mean square error (RMSE) observed for the cognitive measures (ADAS-13 and CDR-SB). While predictions for CSF amyloid-*β* also improved, performance for p-tau showed a slight decline after fine-tuning (a 3.7% increase in RMSE). This divergence likely reflects both differences in the temporal dynamics and the sparsity of observation across biomarkers. Because cognitive assessments are more frequently sampled, fine-tuning tends to prioritise alignment with late-stage clinical decline. Amyloid-*β* accumulates early and typically plateaus before cognitive symptoms, making its trajectory relatively insensitive to these late-stage adjustments. In contrast, p-tau changes most during the transition to cognitive impairment, resulting in a modest reduction in predictive accuracy.

**Table 6.**
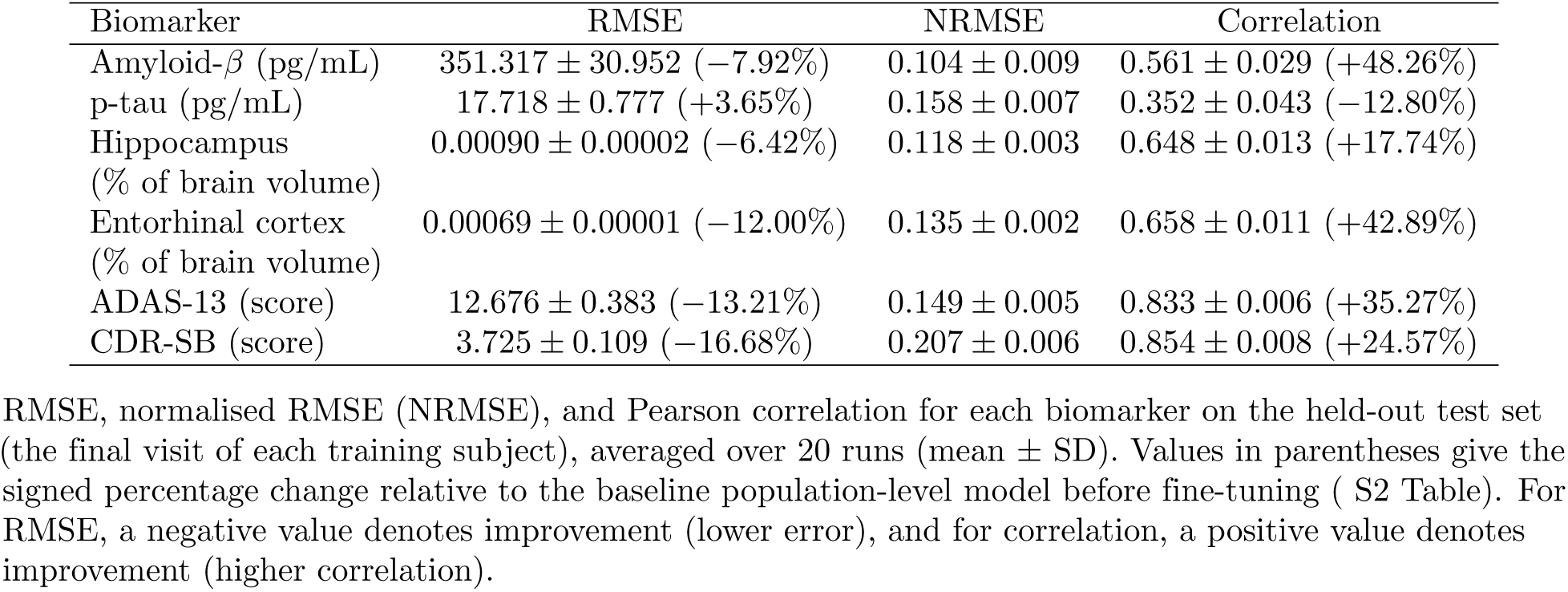
Predictive performance on the held-out test set after subject-specific fine-tuning.

Under the population model, predictive uncertainty was well calibrated across nominal levels (Fig 8a, Table 7, S3 Table). For most biomarkers, empirical coverage tracked the nominal within *±*5% tolerance (Expected Calibration Error (ECE) 2.1-4.8%; Table 7). Amyloid-*β* was the exception (ECE 16.3%), showing over-coverage at all levels (intervals wider than necessary) in both the population and fine-tuned models, consistent with its high estimated observation noise.

**Fig 8.**
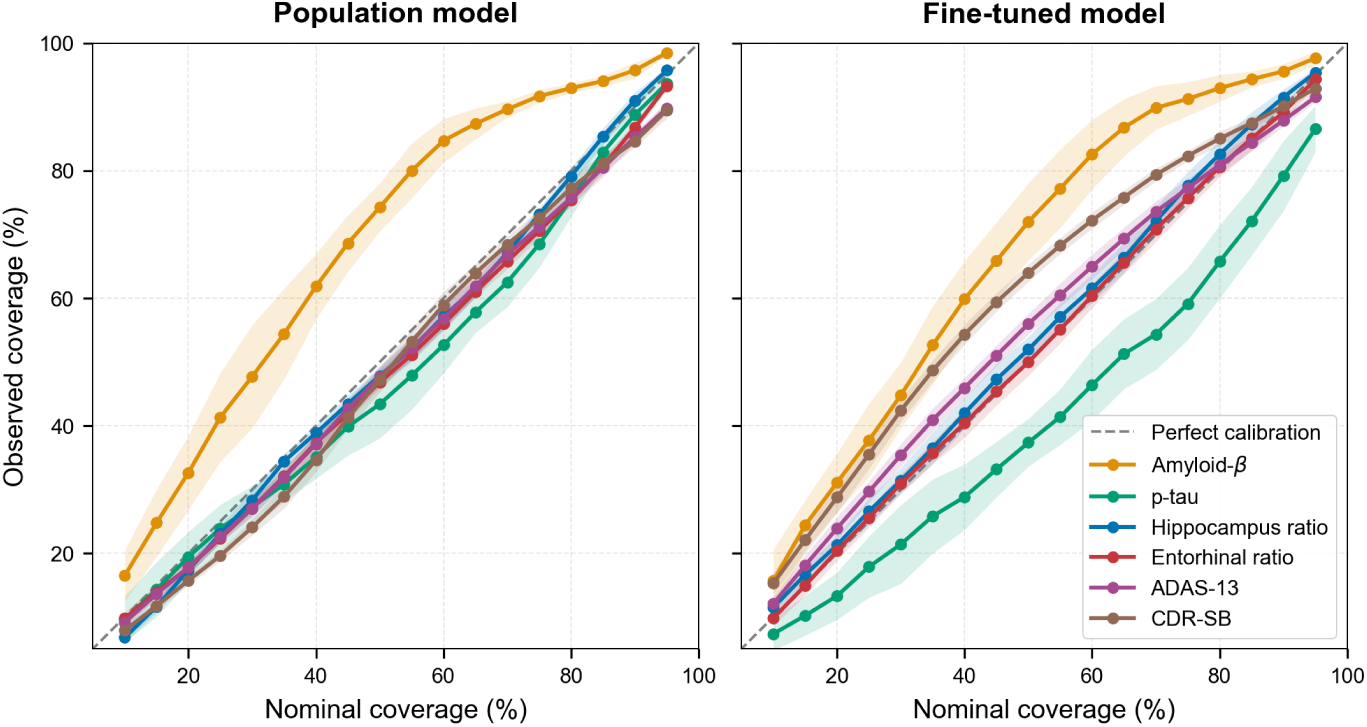
Reliability diagrams for predictive uncertainty on the held-out test set. Observed empirical coverage versus nominal credible level for each biomarker. (**a**) Population-level model. (**b**) After subject-specific fine-tuning. In both panels, the dashed diagonal denotes perfect calibration. Points above the diagonal indicate over-coverage; below indicate under-coverage. Solid lines are the mean coverage across 20 training runs and the shaded bands the *±*1 standard deviation across runs.

**Table 7.** Expected calibration error (ECE) on the held-out test set.

| Biomarker | Population | Fine-tuned | Delta (mean) |
| --- | --- | --- | --- |
| Amyloid-β | $16.27 \pm 2.94\%$ | $14.86 \pm 3.20\%$ | -1.41% |
| p-tau | $4.80 \pm 2.05\%$ | $10.83 \pm 3.68\%$ | +6.03% |
| Hippocampus | $2.09 \pm 0.68\%$ | $2.11 \pm 1.18\%$ | +0.01% |
| Entorhinal cortex | $3.13 \pm 0.57\%$ | $1.47 \pm 0.41\%$ | -1.66% |
| ADAS-13 | $3.13 \pm 0.96\%$ | $4.01 \pm 1.00\%$ | +0.87% |
| CDR-SB | $3.69 \pm 0.91\%$ | $9.09 \pm 0.84\%$ | +5.40% |
| Overall (mean) | $5.52 \pm 4.87\%$ | $7.06 \pm 4.90\%$ | +1.54% |

Mean absolute deviation between observed and nominal coverage, averaged over 18 credible levels from 10% to 95%, for the population-level model and after subject-specific fine-tuning. Lower values indicate better calibration; 0% indicates perfect calibration. The values are averaged across 20 training runs (mean ± SD).

Subject-specific fine-tuning had a mixed, biomarker-dependent effect on calibration (Fig 8b, Table 7). It improved entorhinal cortex (ECE 3.1% *→* 1.5%) and slightly improved amyloid-*β* (16.3% *→* 14.9%, though it remained the worst-calibrated biomarker), while leaving the already well-calibrated hippocampus essentially unchanged (2.1%) and having little effect on ADAS-13 (3.1% *→* 4.0%). However, it degraded calibration for p-tau (4.8% *→* 10.8%) and CDR-SB (3.7% *→* 9.1%). For p-tau, this parallels the decline in its point-prediction accuracy. CDR-SB shows the opposite pattern. Its point predictions improved (Table 6), but its calibration worsened. This likely reflects that fine-tuning was dominated by the frequently sampled

CDR-SB observations. Hence, the model prioritised fitting the CDR-SB mean while holding its predictive variance fixed (only the posterior over *α* and *β* is updated during fine-tuning). This results in more observations falling within the intervals, leading to empirical coverage that overshot the nominal level. Overall, the mean ECE rose from 5.5% to 7.1% across biomarkers, driven by those two.

### Generalisation to unseen subjects

To evaluate generalisation, we assessed predictive performance on a held-out validation cohort of 208 subjects entirely excluded from training (see the Dataset section). For each unseen individual, the progression rate (*α*) and onset time (*β*) were inferred directly from baseline demographic and clinical features (age, sex, APOE*ε*4 status, years of education, and baseline CDR-SB) using the population-trained model, without any fine-tuning. Predictions were then evaluated against that subject’s longitudinal observations.

As detailed in Table 8, the model generalised to unseen subjects using only baseline information. The cognitive measures showed the strongest predictive alignment (ADAS-13: *r* = 0.61 and CDR-SB: *r* = 0.71). Hippocampus volume had the lowest normalised RMSE (0.11). The sparse CSF biomarkers were weakest. P-tau had the lowest correlation (*r* = 0.17) and the highest NRMSE (0.18), and amyloid-*β* also showed only a weak correlation (*r* = 0.37). This likely reflects their greater measurement variability and sparser sampling.

**Table 8.** Predictive performance on the unseen validation cohort.

| Biomarker | RMSE | NRMSE | Correlation | ECE |
| --- | --- | --- | --- | --- |
| Amyloid-β (pg/mL) | 410.101 ± 18.393 | 0.121 ± 0.005 | 0.368 ± 0.037 | 15.07 ± 2.68% |
| p-tau (pg/mL) | 19.627 ± 0.946 | 0.175 ± 0.008 | 0.168 ± 0.016 | 5.81 ± 2.00% |
| Hippocampus<br>(% of brain volume) | 0.00084 ± 0.00001 | 0.109 ± 0.001 | 0.513 ± 0.010 | 4.88 ± 0.68% |
| Entorhinal cortex<br>(% of brain volume) | 0.00064 ± 0.00001 | 0.124 ± 0.002 | 0.438 ± 0.026 | 6.09 ± 0.85% |
| ADAS-13 (score) | 10.890 ± 0.417 | 0.128 ± 0.005 | 0.610 ± 0.035 | 6.99 ± 1.27% |
| CDR-SB (score) | 2.530 ± 0.099 | 0.141 ± 0.006 | 0.710 ± 0.020 | 14.32 ± 1.68% |

Point-prediction accuracy (RMSE, normalised RMSE, Pearson correlation), and level-integrated calibration error (ECE) for each biomarker, evaluated on the held-out validation cohort (subjects excluded from training) using baseline features alone, without fine-tuning. Values are mean ± SD across 20 training runs. ECE is the mean absolute deviation between observed and nominal coverage across 18 credible levels (10-95%), lower being better.

Uncertainty remained reasonably calibrated out-of-sample at the nominal 95% level, where empirical coverage ranged from 89% to 98% across biomarkers (S4 Table). However, calibration was weaker than in-sample. Overall ECE rose to 8.9% (Table 8, Fig 9), compared with 5.5% for the population model and 7.1% after fine-tuning on the held-out test set. Most markers remained moderately calibrated (hippocampus 4.9%, p-tau 5.8%, entorhinal cortex 6.1%, ADAS-13 7.0%), whereas amyloid-*β* (15.1%) and CDR-SB (14.3%) were substantially over-covered across intermediate levels, mirroring their weaker in-sample reliability.

**Fig 9.**
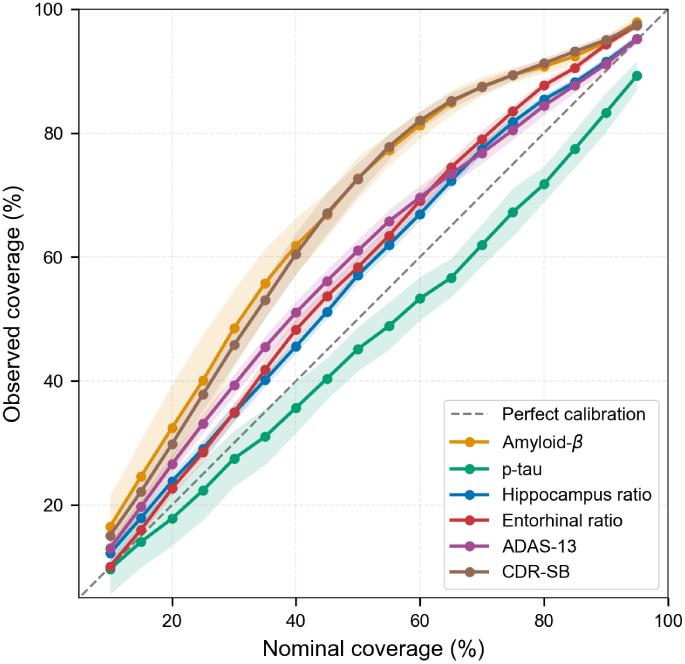
Reliability diagram for predictive uncertainty on the unseen validation cohort. Observed empirical coverage versus nominal credible level for each biomarker, evaluated on entirely unseen subjects from baseline features alone without fine-tuning. The dashed diagonal denotes perfect calibration. Points above the diagonal indicate over-coverage; below indicate under-coverage. Solid lines are the mean coverage across 20 training runs and the shaded bands the *±*1 standard deviation across runs.

## Discussion

In this study, we introduce a variational disease progression score (DPS) framework to model the biomarker dynamics of Alzheimer’s disease from sparse, multimodal cohort data. By estimating subject-specific onset times (*β*) and progression rates (*α*), the model aligns heterogeneous individuals onto a continuous latent disease timeline. The variational formulation further enables subject-level uncertainty quantification, providing predictive intervals that are well calibrated for most biomarkers, and bridging population-level inference with personalised prognosis.

Clinically, the cascade strengths estimated within the imposed topology are consistent with current models of Alzheimer’s disease progression, including the amyloid cascade hypothesis [9]. Among the pre-specified links, the strongest estimated strengths connected tau pathology to medial temporal atrophy, while the amyloid-*β →* p-tau link was the weakest but most reproducibly estimated, which is consistent with amyloid acting as a slow upstream initiator rather than an immediate driver. The model also recovered established biology. It learned mappings from baseline features to progression parameters that reproduced the expected effect of APOE*ε*4 genotype groups on onset and rate. Lastly, without diagnostic labels as inputs, the inferred DPS correctly separated the cognitively normal, MCI, and AD groups, an emergent alignment with clinical staging.

A key strength of this framework lies in its utility as a dynamic clinical prognosis tool. By combining the interpretability of traditional DPS models with the scalability of probabilistic variational inference, it extends beyond static point estimation to provide individualised trajectory prediction with quantified uncertainty. The model produces baseline prognostic estimates for unseen individuals from demographic and clinical features, with accuracy that varies by biomarker, strongest for the cognitive measures, and these can be dynamically fine-tuned as longitudinal data accrue. Fine-tuning reduced parameter uncertainty and improved point-prediction accuracy for most biomarkers, although its effect on interval calibration was mixed.

Despite these advances, several limitations should be considered. First, reliance on sparse and heterogeneous longitudinal data necessitates simplifying assumptions regarding disease dynamics. To maintain interpretability and tractability, the current framework assumes logistic-like biomarker trajectories along a single latent disease axis, in alignment with the theoretical model proposed by Jack et al. [9]. Consequently, this approach inherently disregards heterogeneity and the potential existence of distinct disease subtypes. In addition, the model currently restricts structural neuroimaging to only two regions, the hippocampus and the entorhinal cortex, chosen for their proximity and shared role in early memory impairment. This restricts the understanding of broader cortical disease spread and potential temporal lags between region-specific biomarker changes. This is consistent with our findings that pathways linking medial temporal lobe atrophy to CDR-SB showed weaker statistical support, likely reflecting the broader functional domains captured by this measure. This suggests that extending the framework to incorporate distributed cortical regions or more granular clinical assessment may improve its ability to capture the full spectrum of disease progression.

Second, the imbalance in observation frequency affects subject-specific fine-tuning. Because cognitive measures are sampled far more frequently than fluid biomarkers, the model may prioritise alignment with late-stage clinical decline, potentially leading to minor temporal misalignments in more dynamic intermediate biomarkers such as p-tau. This also introduces a calibration trade-off: point accuracy improved, while the fixed-width intervals became miscalibrated for CDR-SB and p-tau. Because the observation noise is not updated during fine-tuning, gains in mean accuracy are not always matched by appropriate interval widths.

From a methodological perspective, the use of variational inference enables scalability to large, multimodal datasets but may underestimate posterior uncertainty compared to fully Bayesian approaches such as Markov Chain Monte Carlo (MCMC) [46]. In addition, although the framework demonstrated robust performance on held-out temporal visits and entirely unseen subjects within the study cohort, its reliability on out-of-distribution datasets and its capacity for long-term extrapolation beyond the observed follow-up window remain to be established through external, real-world longitudinal validation.

## Conclusion

We presented a variational Disease Progression Score framework that places Alzheimer’s disease biomarker trajectories on a common latent timeline while quantifying subject-level uncertainty. On the ADNI cohort, the model produced individualised, uncertainty-aware prognoses from baseline features, recovered biomarker orderings and cascade strengths consistent with established disease biology, and generalised to entirely unseen subjects. Its uncertainty estimates were well calibrated for most biomarkers, though calibration degraded for amyloid-*β* and CDR-SB. Its reliability on out-of-distribution data and its ability to forecast beyond the follow-up intervals available in this study both remain to be established. By combining the interpretability of disease progression modelling with calibrated, updatable uncertainty, the framework offers a step towards probabilistic, individualised prognosis in Alzheimer’s disease.

## Data Availability

The data used in this study were obtained from the Alzheimer's Disease Neuroimaging Initiative (ADNI) database (https://adni.loni.usc.edu).

## Supporting information

**S1 Fig. Training loss convergence across 20 independent runs.** Median training losses on a log-scale y-axis across epochs, with 68% and 95% interval bands over the 20 runs. E-step reconstruction loss (*L^NLL^*, upper left), E-step KL loss (*L^KL^*, upper right), M-step loss (*L_M_*, lower left), and total loss (*L_E_* + *L_M_*, lower right). The reconstruction and cascade (M-step) losses are negative log-likelihoods, with lower values indicating a better fit. For the KL term, values closer to zero indicate posteriors closer to their priors.

**S1 Table. Best training losses across 20 independent runs.** Mean *±* standard deviation of the best loss achieved per run for each objective component. The reconstruction and cascade losses are negative log-likelihoods, with lower values indicating a better fit. For the KL term, values closer to zero indicate posteriors closer to their priors.

**S2 Fig. Predictive performance on the held-out test set after subject-specific fine-tuning.** Scatter plots comparing model-predicted versus observed biomarker values in the held-out test set after subject-specific fine-tuning. Each data point corresponds to an individual visit, coloured by diagnostic group (cognitively normal, CN; mild cognitive impairment, MCI; and Alzheimer’s disease, AD). The dashed grey diagonal represents the perfect prediction. Performance metrics, including Pearson correlation (*r*), coefficient of determination (*R*^2^), root mean square error (RMSE), and the total number of observations (*n*), are shown in the inset for each biomarker.

**S2 Table. Predictive performance on the held-out test set for the baseline population-level model.** RMSE, normalised RMSE (NRMSE), and Pearson correlation for each biomarker on the held-out test set (the final visit of each training subject), averaged over 20 runs (mean ± SD).

**S3 Fig. Learned aleatoric observation noise across biomarker modalities.** The inferred standard deviation (*σ*) of the modality-specific observation noise is plotted as a function of the latent Disease Progression Score (DPS). Fluid biomarkers (amyloid-*β*, p-tau) and structural neuroimaging (hippocampus and entorhinal cortex) were modelled as homoscedastic, yielding a constant noise throughout the disease course. The cognitive measurements (ADAS-13 and CDR-SB) were modelled as heteroscedastic, yielding an exponential increase as the disease advanced.

**S3 Table. Empirical coverage of 95% credible intervals on the held-out test set.** Percentage of held-out test observations (the final visit of each training subject) falling within the predicted 95% credible intervals. Values are compared between results from the baseline population-level model and after personalised fine-tuning. A well-calibrated model yields coverage near 95%. Values above indicate over-coverage, and values below indicate under-coverage.

**S4 Table. Empirical coverage of 95% credible intervals on the unseen validation cohort.** Percentage of observations from the unseen validation cohort falling within the predicted 95% credible intervals. A well-calibrated model yields coverage near 95%. Values above indicate over-coverage, and below indicate under-coverage.

## Acknowledgments

We thank Professor Christopher Kipps and Dr Christine Evers for helpful comments on the work.

